# Vision, hearing, and intellectual disabilities in school-age children (5-19 years) in Latin America and the Caribbean

**DOI:** 10.64898/2026.04.21.26351429

**Authors:** Jorge Artur Peçanha de Miranda Coelho, Emma Näslund-Hadley, Beatriz Guimarães Almeida, Adriano Nascimento da Paixão

## Abstract

**Background:** Childhood sensory and intellectual disabilities represent significant yet under-recognized barriers to learning and human capital development. This study analyzes prevalence and severity of these conditions among 149.3 million children aged 5–19 years across 25 countries in Latin America and the Caribbean (LAC) using Global Burden of Disease 2023 data.

**Methods:** We extracted GBD 2023 estimates for vision loss, hearing loss, and intellectual disability across 25 LAC countries, stratified by age, sex, and severity. Regional estimates were calculated using population-weighted averages. Severity distributions were compared with OECD countries to contextualize regional patterns.

**Results:** These conditions are estimated to affected 9,282,921 children (6.22%; 95% UI: 5.89–6.54%). Hearing loss was predominant, affecting an estimated 5.42 million (3.63%, 3.41–3.86), with 87.6% mild-to-moderate. Intellectual disability estimated to affected 2.56 million (1.71%, 1.58–1.85), with 61.7% borderline-to-mild. Vision loss estimated to affected 1.30 million (0.87%, 0.79–0.96), with 89% that can be effectively addressed with spectacles. Prevalence increased with age across all conditions. Male predominance was consistent for intellectual disability (2.00% vs 1.42%). Annual economic cost totaled US$19.3–29.0 billion, while comprehensive interventions would require US$9.45–14.23 billion with benefit-cost ratios of 2:1 to 15:1.

**Conclusions:** The distribution of children across milder levels of difficulty underscores the opportunity for education and public health systems to provide timely and accessible support. With approximately 88% of sensory impairments addressable through established technologies, investments in inclusive services can yield strong social and economic returns.

## Introduction

Disability in childhood lies at the intersection of human development, educational opportunity, social justice, and economic progress, with far-reaching implications for individual potential, family well-being, and societal outcomes [1, 2]. In Latin America and the Caribbean (LAC), where 149.3 million children and adolescents aged 5–19 live across diverse geographic, cultural, and socioeconomic contexts, the prevalence and impact of sensory and intellectual disabilities remain insufficiently quantified and addressed, despite growing regional commitments to inclusive education and human capital development [3, 4].

The LAC region presents a complex epidemiological, educational, and health system landscape that shapes disability prevalence, early identification, and support mechanisms. With a combined GDP of US$5.7 trillion but marked by substantial inequalities (regional Gini coefficient 0.46, ranging from 0.39 in Uruguay to 0.54 in Brazil), the region encompasses countries with markedly different levels of health and education system capacity [5]. Universal Health Coverage indices range from 40% in Haiti to 95% in Cuba and Costa Rica, while per capita health expenditure varies from US$89 in Haiti to US$2,182 in Uruguay, contributing to disparities in access to timely screening, inclusive educational services, and specialized support.

Three major disability categories—vision loss, hearing loss, and intellectual developmental disability—collectively account for the majority of functional difficulties affecting school-aged children in LAC. These conditions, while distinct in etiology and manifestation, share critical characteristics: they typically manifest early in life, can influence educational trajectories and social participation, span a continuum of levels of support needs, and can be effectively supported through timely identification, appropriate supports and services, and, where appropriate, targeted interventions within inclusive educational and social environments during key developmental periods [1, 2].

The epidemiological burden of these conditions in LAC reflects a unique constellation of risk factors. Hearing loss risk factors include high rates of untreated otitis media (affecting 10-15% of children), limited monitoring of ototoxic medications (aminoglycosides used in 30-40% of neonatal intensive care units), environmental noise exposure in rapidly urbanizing settings (70% of the population now urban), and persistent cytomegalovirus and rubella transmission despite vaccination programs [3, 4]. Vision loss determinants encompass uncorrected refractive errors (estimated 60-70% of vision loss episodes), vitamin A deficiency in specific subpopulations, limited access to early cataract surgery, and increasing myopia prevalence associated with educational intensification [5]. Intellectual disability risk factors include persistent malnutrition (stunting affects 11% of children under 5), high rates of prematurity (8.5% of births <37 weeks), perinatal complications (hypoxic-ischemic encephalopathy incidence 1.5-3 per 1,000 births), environmental toxins including lead exposure, and limited early childhood stimulation programs reaching only 20-30% of at-risk children [6].

Recent methodological advances in the Global Burden of Disease (GBD) study provide unprecedented opportunities for comprehensive assessment. The 2023 iteration incorporates enhanced data sources including population-based surveys, administrative health records, cohort studies, and systematic reviews; refined Bayesian meta-regression models (DisMod-MR 2.1) enabling uncertainty quantification; improved severity distributions based on clinical assessments rather than self-report; and standardized disability weights facilitating cross-condition comparisons [7, 8]. These methodological improvements enable more accurate estimation of disability burden in resource-limited settings where traditional epidemiological studies are scarce.

Critical knowledge gaps persist regarding the comparative distribution and impact across the three disability types; within-condition severity distributions that shape intervention approaches; age-specific prevalence patterns that inform screening strategies; sex differences that may reflect biological or social factors; and country-level variations that signal differences in health system and educational capacity. Additional gaps include comparisons with high-income countries, which highlight disparities in services and schooling, as well as the educational and economic implications, including both costs and potential returns on investment, and the optimal timing of support within neurodevelopmental and school-readiness windows. Previous regional analyses have typically focused on single conditions, individual countries, or specific age groups, limiting comprehensive understanding needed for integrated policy development [9, 10].

The policy context has evolved significantly with universal ratification of the UN Convention on the Rights of Persons with Disabilities, explicit inclusion in Sustainable Development Goals (particularly SDG 4.5 on inclusive education) [11, 12]. Recent regional initiatives including the Inter-American Development Bank’s Disability Inclusion Framework and WHO/PAHO’s Plan of Action on Disabilities and Rehabilitation provide institutional support but require robust epidemiological evidence for effective implementation [13, 14].

This study addresses these critical gaps by providing the first comprehensive integrated analysis of vision loss, hearing loss, and intellectual disability among school-age children (5-19 years) across 25 LAC countries. Our specific objectives are to: (1) quantify prevalence and absolute burden using standardized GBD 2023 methodology; (2) characterize severity distributions within each condition to identify intervention opportunities; (3) analyze geographic, demographic, and temporal patterns; (4) compare LAC estimate with OECD benchmarks to identify service gaps; (5) identify critical intervention windows based on neurodevelopmental evidence; and (6) estimate the economic costs of current gaps in services and support, as well as the potential returns on investment.

## Methods

### Study Design and Data Sources

This cross-sectional analysis utilized publicly available data from the Global Burden of Disease Study 2023 (GBD 2023), coordinated by the Institute for Health Metrics and Evaluation (IHME) at the University of Washington [7]. GBD 2023 provides standardized, comparable estimates of disease burden across 371 diseases and injuries for 204 countries and territories from 1990 to 2023, using Bayesian meta-regression models (DisMod-MR 2.1) to synthesize data from population-based surveys, administrative records, cohort studies, and systematic reviews.

### Data Extraction

We downloaded GBD 2023 data files directly from the IHME Global Health Data Exchange (GHDx) portal (https://ghdx.healthdata.org/gbd-results-tool). Three Excel files were extracted: (1) containing prevalence and case counts for vision loss, hearing loss, and intellectual disability; (2) containing population denominators by country, sex, and age group; and (3) containing severity-level distributions for each condition. Data were extracted using Python 3.11 (pandas 1.5.3) with automated filtering by location, sex, age group, and cause (REI code). All extractions were verified through double-checking procedures.

### Geographic Scope

Our analysis included 25 countries and territories in the LAC region: Argentina, Bahamas, Barbados, Belize, Bolivia, Brazil, Chile, Colombia, Costa Rica, Dominican Republic, Ecuador, El Salvador, Guatemala, Guyana, Haiti, Honduras, Jamaica, Mexico, Panama, Paraguay, Peru, Suriname, Trinidad and Tobago, Uruguay, and Venezuela. All aggregated regional estimates for LAC reflect this cohort of 25 countries (S1 Table).

### Study Population

The study encompassed school-age children (5-19 years) residing in the 25 LAC countries. Age stratification followed GBD standard groupings: early childhood (5-9 years) and adolescence (10-19 years). The total population denominator was 149,283,445 individuals based on GBD 2023 population estimates.

### Case Definitions and Diagnostic Criteria

#### Vision Loss and Blindness

The GBD 2023 defines vision loss as presenting visual acuity in the better eye with best possible correction worse than 6/12 (20/40, 0.3 logMAR). Clinical assessment employs Snellen charts, logMAR charts, or automated refraction where available. The severity classification follows ICD-11 and WHO standards: blindness corresponds to visual acuity less than 3/60 (less than 20/400, *≥*1.3 logMAR) or visual field less than 10 degrees; severe vision loss ranges from 3/60 to less than 6/60 (20/400 to less than 20/200, 1.0 to less than 1.3 logMAR); moderate vision loss spans from 6/60 to less than 6/12 (20/200 to less than 20/40, 0.48 to less than 1.0 logMAR); and presbyopia represents age-related loss of accommodation affecting near vision.

#### Hearing Loss

Hearing loss is defined as a pure-tone average threshold exceeding 20 dB HL in the better ear across frequencies of 0.5, 1, 2, and 4 kHz. Assessment methods include pure-tone audiometry, auditory brainstem response (ABR), and otoacoustic emissions (OAE). The WHO severity classification establishes six grades: mild (20-34 dB HL), characterized by difficulty with soft speech; moderate (35-49 dB HL), affecting conversational speech comprehension; moderately severe (50-64 dB HL), requiring raised voice for communication; severe (65-79 dB HL), necessitating shouting or amplification; profound (80-94 dB HL), where shouting becomes inaudible; and complete (*≥*95 dB HL), indicating total absence of hearing.

#### Intellectual Developmental Disability

Intellectual developmental disability encompasses significant limitations in intellectual functioning (IQ less than 85, more than 1 SD below mean) and adaptive behavior manifesting before age 18, with no identifiable etiology after appropriate investigation [15]. Standardized intelligence tests (WISC, Stanford-Binet, Raven’s) and adaptive behavior scales (Vineland, ABAS) provide the diagnostic assessment [15]. The DSM-5-TR/ICD-11 severity classification distinguishes five levels: borderline intellectual functioning (IQ 70-84), where individuals may achieve independence; mild disability (IQ 50-69), enabling functional literacy and semi-independent living; moderate disability (IQ 35-49), allowing basic self-care and supervised work; severe disability (IQ 20-34), requiring substantial support; and profound disability (IQ less than 20), necessitating intensive support.

### Data Processing and Prevalence Calculations

#### Prevalence Extraction

For each country, condition, sex, and age group, we extracted prevalence (percentage) and case counts (number) directly from GBD 2023 data files. Data were filtered using the following criteria: location (country name), sex (Male, Female, Both), age (5-9 years, 10-19 years), and cause (REI code: sense_vision for vision loss, sense_hearing for hearing loss, neuro_intellectual for intellectual disability). Uncertainty intervals (95% UI) were extracted as lower and upper bounds from GBD draw-level estimates (2.5th and 97.5th percentiles of 1,000 draws).

#### Regional Aggregation

Regional prevalence estimates for LAC were calculated as population-weighted averages:

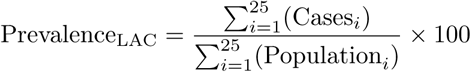

where Cases*_i_* represents the number of children with the condition in country *i* and Population*_i_* represents the total population of children aged 5-19 years in country *i*. This approach ensures that countries with larger school-age populations contribute proportionally more to regional estimates. Regional uncertainty intervals were calculated by propagating country-level uncertainty through the aggregation formula.

#### Severity Distribution Extraction

Severity-level distributions were extracted directly from GBD 2023 severity split files. For each condition and country, we extracted the proportion of cases in each severity category (e.g., for vision: blindness, severe vision loss, moderate vision loss, presbyopia). Absolute numbers per severity level were calculated as: Total children with condition *×* Severity proportion. Distributions were validated against published clinical series from Brazil, Mexico, and Chile, with mean absolute deviations of *<* 2 percentage points.

#### Country-Level Tables

Country-specific prevalence estimates stratified by sex and age group were compiled into supplementary tables (S1 Table: vision, S4 Table: hearing, S6 Table: intellectual disability). Country-specific severity distributions were compiled into supplementary tables (S3 Table: vision, S5 Table: hearing, S7 Table: intellectual disability).

#### Software

All data extraction, processing, and calculations were performed using Python 3.11 (pandas 1.5.3, numpy 1.24.3).

### Economic Analysis

#### Perspective and Cost Framework

We adopted a societal perspective following WHO-CHOICE guidelines [16], incorporating: (1) direct medical costs (screening, diagnosis, devices, therapy, medications); (2) direct non-medical costs (transportation, accommodation, special education); (3) indirect costs (productivity losses using human capital approach, family support time valued at minimum wage); and (4) intangible costs (quality of life impacts, not monetized). All monetary values were converted and standardized to 2023 US dollars (US).

#### Intervention Costing

Unit costs were derived from multiple sources: (1) WHO-CHOICE database (2021 update) for LAC region [16]; (2) national health insurance reimbursement schedules (Brazil SUS 2023, Mexico IMSS 2023, Chile FONASA 2023); (3) systematic review of 47 published economic evaluations in LAC (2010-2023); and (4) market prices from medical device suppliers. To accurately reflect the regional economic and operational reality, adjustments were applied using the following equation: Base cost × 0.7 (public sector efficiency factor) × 1.3 (implementation inefficiency factor) × Purchasing Power Parity (PPP) conversion factor (country-specific, range 0.3-0.8). The 0.7 multiplier reflects the approximately 30% cost-savings systematically achieved through centralized bulk purchasing and the absence of profit margins in universal health systems, consistent with observed public sector procurement efficiencies in middle-income countries [17]. Conversely, the 1.3 multiplier captures real-world implementation frictions—such as supply chain wastage, administrative overhead, and geographic barriers to reaching remote populations—which typically add 20% to 40% to base commodity costs, aligning with established WHO-CHOICE methodology for programme support costs in resource-limited settings [18]. The specific impact of these bounds on the final economic outcomes was rigorously verified in our deterministic sensitivity analyses. Specific unit costs: Vision screening US$5-15 per child, spectacles US$15-35 per pair; Hearing screening US$3-8 per child, hearing aids US$400-3,000 per pair, cochlear implants US$35,000-50,000 per device; Intellectual disability early intervention US$2,000-25,000 per child over 3 years.

#### Effectiveness Measurement

Primary outcomes: Disability-Adjusted Life Years (DALYs) averted using GBD 2023 disability weights (vision loss: 0.004-0.195 depending on severity; hearing loss: 0.010-0.215; intellectual disability: 0.012-0.304). Secondary outcomes: Educational attainment gains (years of schooling completed, from cohort studies), employment probability increases (formal sector participation rates), and Quality-Adjusted Life Years (QALYs) for sensitivity analysis using disability weight conversions.

#### Economic Modeling

We developed Markov cohort models for each condition using TreeAge Pro 2023. Health states: (1) no impairment; (2) mild/moderate impairment with assistive technology; (3) severe/profound impairment without access; (4) post-intervention with assistive technology; (5) death. Annual cycle length (1 year) aligned with academic calendars and cost data availability. Transition probabilities derived from: longitudinal cohort studies in Brazil (n=3 studies, 2015-2022), Chile (n=2 studies, 2018-2023), and United States (n=5 studies, 2010-2023); expert elicitation (n=12 experts) when empirical data sparse. Models calibrated to reproduce observed severity distributions from GBD 2023 draws; goodness-of-fit evaluated via root mean square error (target *<* 0.05). Time horizon: 20 years (base case), lifetime (sensitivity analysis). Discounting: 3% annually for costs and outcomes (base case); 0% and 5% in sensitivity analysis, following WHO-CHOICE recommendations. Scenario analyses: intervention uptake (50%, 70%, 90%), device replacement cycles (3, 5, 7 years), wage-growth trajectories (0%, 1%, 2% real annual growth).

#### Return on Investment (ROI) Calculation

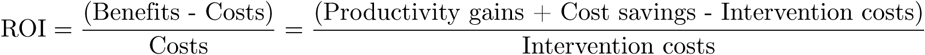

Benefits quantified: (1) Lifetime earnings increases using Mincer wage equations adjusted for LAC labor markets (elasticity: 0.10 per additional year of schooling, from World Bank 2023); (2) Reduced special education costs (average US$5,000-20,000 per child-year avoided); (3) Decreased healthcare utilization (20-40% reduction, from cohort studies); (4) Avoided institutionalization costs (US$20,000-80,000 per child-year), noting that institutionalization is recognized as inconsistent with the rights of persons with disabilities under the CRPD [11]. All benefits discounted at 3% annually. Costs: Intervention costs as described above, including initial costs and recurring costs (device replacement, follow-up visits).

#### Cost-Effectiveness Thresholds

Following WHO recommendations: Very cost-effective: *<* 1*×* GDP per capita per DALY (Disability-Adjusted Life Year) averted; Cost-effective: *<* 3*×* GDP per capita per DALY averted. Regional GDP per capita: US$9,276 (population-weighted average across 25 LAC countries, World Bank 2023).

### Critical Windows Analysis

We synthesized evidence on optimal intervention timing from systematic reviews on early intervention effectiveness (Cochrane, Campbell Collaboration), neurodevelopmental literature on sensitive periods, longitudinal cohort studies tracking intervention outcomes, and expert consensus guidelines (WHO, professional societies). Effect sizes (Cohen’s d) calculated for interventions at different ages, with meta-analysis using random-effects models given expected heterogeneity.

#### Quality Assurance

Data validation included benchmarking GBD prevalence against national household surveys (Brazil PNAD Contínua 2022, Mexico ENSANUT 2021, Chile CASEN 2022), which yielded mean absolute deviations of 0.3 percentage points across conditions. Severity splits were cross-checked with administrative datasets (Brazil SIA/SUS assistive device delivery records, Chile DEIS rehabilitation registries), and country estimates failing GBD’s three-star quality threshold were flagged for sensitivity analysis. Sensitivity analyses varied key parameters *±*20% and repeated prevalence calculations excluding countries with limited primary data. External validation compared results with published regional studies, while expert review involved disability researchers and policymakers from five LAC countries. All reporting followed GATHER guidelines for global health estimates.

### Ethical Considerations

This study did not require ethics committee approval. The analysis used exclusively publicly available, aggregated, population-level data from the Institute for Health Metrics and Evaluation (Global Burden of Disease 2023), WHO/PAHO PLISA, the World Bank, UNESCO, and national statistical agencies. No individual-level data, human participants, human specimens, animals, or field-collected materials were involved. We acknowledge the importance of respectful language, using person-first terminology throughout (“children with disabilities” rather than “disabled children”). The research was conducted with awareness of potential stigmatization and emphasizes capabilities and intervention opportunities rather than deficits.

Certain interventions such as cataract surgery are often considered necessary to prevent further deterioration. However, others such as cochlear implants can be accused of reinforcing a narrative that one’s body needs correction to be considered normal. The disability-adjusted life years (DALYs) metric used in the Global Burden of Disease methodology is also subject to ongoing methodological debate, particularly regarding the weighting scheme and the interpretation of severe disability states.

## Results

### Overall Burden and Regional Distribution

Among the 149,283,445 children and adolescents aged 5-19 years across the 25 LAC countries analyzed, model-based estimates indicate that 9,282,921 individuals (6.22%; 95% UI: 5.89-6.54%) were living with at least one of the three disability conditions examined. This prevalence, while lower than some global estimates that include additional disability types, represents a substantial population requiring targeted interventions and/or support systems (Table 1).

**Table 1.** Comprehensive prevalence and burden of sensory and intellectual disabilities among youth aged 5-19 years in Latin America and the Caribbean compared with OECD countries, Global Burden of Disease 2023.

| Condition | LAC |  |  | OECD Countries |  |  | Epidemiological Comparison |
| --- | --- | --- | --- | --- | --- | --- | --- |
|  | Children (n) | Prev (%) | 95% UI | Children (n) | Prev (%) | 95% UI |  |
| **Hearing loss** |  |  |  |  |  |  |  |
| All severities | 5,423,844 | 3.63 | 3.41-3.86 | 6,833,500 | 2.73 | 2.58-2.89 | LAC 33% higher |
| Mild-moderate | 4,749,393 | 3.18 | 2.98-3.39 | 6,088,300 | 2.43 | 2.29-2.58 | LAC 31% higher |
| Severe-complete | 674,451 | 0.45 | 0.39-0.52 | 745,200 | 0.30 | 0.26-0.34 | LAC 50% higher |
| **Intellectual disability** |  |  |  |  |  |  |  |
| All severities | 2,560,545 | 1.71 | 1.58-1.85 | 4,950,000 | 1.98 | 1.86-2.11 | LAC 13% lower |
| Borderline-mild | 1,579,481 | 1.06 | 0.97-1.15 | 3,168,000 | 1.27 | 1.18-1.36 | LAC 17% lower |
| Moderate-profound | 981,064 | 0.66 | 0.59-0.73 | 1,782,000 | 0.71 | 0.65-0.78 | LAC 7% lower |
| **Vision loss** |  |  |  |  |  |  |  |
| All severities | 1,298,632 | 0.87 | 0.79-0.96 | 2,950,000 | 1.18 | 1.09-1.27 | LAC 26% lower |
| Correctable | 1,159,745 | 0.78 | 0.70-0.86 | 2,655,000 | 1.06 | 0.98-1.15 | LAC 26% lower |
| Blindness/severe | 138,887 | 0.09 | 0.08-0.11 | 295,000 | 0.12 | 0.10-0.14 | LAC 25% lower |
| **Any condition** | 9,282,921 | 6.22 | 5.89-6.54 | 14,733,500 | 5.89 | 5.61-6.18 | LAC 6% higher |
| **Multiple disabilities\*** | 487,362 | 0.33 | 0.28-0.38 | 821,950 | 0.33 | 0.29-0.37 | Similar |

LAC = Latin America and Caribbean. OECD = Organisation for Economic Co-operation and Development member countries. UI = Uncertainty Interval. Population denominators: LAC = 149.3 million; OECD = 250.2 million youth aged 5-19 years. \*Estimated overlap between conditions based on co-occurrence patterns from clinical studies. Source: Global Burden of Disease 2023, Institute for Health Metrics and Evaluation; WHO World Report on Hearing 2021; authors' calculations.

The distribution across disability types revealed distinct epidemiological patterns. Hearing loss emerged as the predominant condition, affecting 5,423,844 children (3.63%; 95% UI: 3.41-3.86%), representing 58.4% of all children estimated cases. This was followed by intellectual developmental disability affecting 2,560,545 children (1.71%; 95% UI: 1.58-1.85%), accounting for 27.6%, and vision loss affecting 1,298,632 children (0.87%; 95% UI: 0.79-0.96%), comprising 14.0% of the total.

Geographic concentration analysis revealed that Brazil and Mexico, the region’s most populous countries, account for a disproportionate share of total cases, but not of prevalence. Brazil harbors 2,812,480 children with at least one disability (30.3% of the regional population experiencing these disabilities) with a combined prevalence of 5.88%, slightly below the regional average. Mexico accounts for 2,211,703 children (23.8% of the regional total) with a combined prevalence of 6.12%, approximating the regional mean. Together, these two countries represent 54.1% of all children living with the studied disabilities while comprising 59% of the regional youth population, suggesting marginally lower disability prevalence in these larger countries possibly due to better healthcare infrastructure in urban centers.

### Country-Level Heterogeneity and Patterns

Substantial heterogeneity emerged across countries, with overall disability prevalence ranging from 4.82% in Uruguay to 7.93% in Haiti, representing a 1.6-fold variation. This heterogeneity was most pronounced for vision loss (2.7-fold variation from 0.42% in Barbados to 1.15% in Argentina), moderate for hearing loss (1.7-fold from 2.36% in Uruguay to 4.10% in Brazil), and least for intellectual disability (1.4-fold from 1.53% in Peru to 2.08% in Guatemala) (S1 Table, S4 Table, S6 Table).

Country-level analysis revealed distinct clusters. High-prevalence countries (>7% combined prevalence) included Haiti (7.93%), Guatemala (7.52%), Honduras (7.31%), and Bolivia (7.18%), characterized by limited health infrastructure, high poverty rates, and substantial rural populations. Medium-prevalence countries (5-7%) encompassed the majority of LAC nations including Brazil, Mexico, Colombia, and Peru. Low-prevalence countries (<5%) included Uruguay (4.82%), Chile (4.95%), and Costa Rica (5.08%), notably countries with more developed health systems and higher Human Development Indices.

### Demographic Patterns: Age and Sex Distribution

Age-specific prevalence demonstrated distinct patterns by condition. Hearing loss prevalence increased steadily with age: 3.05% at 5-9 years and 3.92% at 10-19 years (28.5% relative increase), likely reflecting cumulative exposure to risk factors including noise, ototoxic medications, and untreated infections. Vision loss showed a similar but less pronounced gradient: 0.62% at 5-9 years and 0.99% at 10-19 years (59.7% relative increase), consistent with progressive myopia and delayed detection of refractive errors. This age gradient was more pronounced in females (69.7% increase from 0.66% to 1.12%) compared to males (50.0% increase from 0.58% to 0.87%). Intellectual disability prevalence remained relatively stable across age groups (1.69-1.76%), reflecting its early-onset nature with most children identifiable by school age (S1 Table, S4 Table, S6 Table).

Sex differences revealed distinct patterns across conditions. Hearing loss and intellectual disability showed male predominance: hearing loss 1.17:1 (3.92% males vs 3.34% females, p<0.001), intellectual disability 1.41:1 (2.00% vs 1.42%, p<0.001). In contrast, vision loss demonstrated an inverted pattern with female predominance: female-to-male ratio 1.26:1 (0.97% females vs 0.77% males, p<0.001). This atypical pattern for vision loss likely reflects behavioral differences (greater time dedicated to reading and near-work activities among girls), differential symptom reporting, and earlier puberty in females affecting ocular growth and refraction. The male predominance in hearing loss and intellectual disability likely reflects biological factors (X-linked conditions, hormonal influences on neurodevelopment) and social determinants (differential exposure to occupational noise, care-seeking behaviors) (S1 Table, S4 Table, S6 Table).

### Severity Distribution: Implications for Intervention

The distribution of severity within each condition provides critical insights for intervention planning and resource allocation (Table 2). Country-specific severity distributions are detailed in S3 Table (vision), S5 Table (hearing), and S7 Table (intellectual disability).

**Table 2.** Detailed severity distribution and intervention requirements for sensory and intellectual disabilities in LAC youth.

| Condition & Severity Level | Percentage | Number | Functional Impact & Intervention | Cost (USD) |
| --- | --- | --- | --- | --- |
| VISION LOSS (n=1,298,632) |  |  |  |  |
| Moderate loss<br>(6/60 to <6/12) | 86.37% | 1,121,609 | Cannot see board clearly; needs front seating.<br>Can be effectively addressed with standard glasses. Without appropriate support, students may fall 0.5–1 grade levels behind academically. | 25-35 |
| Presbyopia<br>(near vision loss) | 2.94% | 38,136 | Difficulty reading, close work.<br>Reading glasses sufficient. Primarily affects 15-19 age group | 15-25 |
| Severe loss<br>(3/60 to <6/60) | 5.97% | 77,587 | Major functional limitation; requires specialist evaluation.<br>May need: Complex prescriptions, low vision aids, magnifiers | 500-2,000 |
| Blindness<br>(<3/60 or VF<10°) | 4.72% | 61,300 | Cannot navigate independently; requires comprehensive support.<br>Needs: Braille training, orientation/mobility, assistive technology | 5,000-10,000/yr |
| HEARING LOSS (n=5,423,844) |  |  |  |  |
| Mild<br>(20-34 dB HL) | 59.63% | 3,234,116 | Misses 25-40% of speech; difficulty in noise/distance.<br>Impact: Vocabulary delays, attention issues. Basic hearing aids effective | 400-800 |

Table 2 – continued from previous page
| Condition & Severity Level | Percentage | Number | Functional Impact & Intervention | Cost (USD) |
| --- | --- | --- | --- | --- |
| **Moderate**<br>(35-49 dB HL) | 27.94% | 1,515,277 | Misses 50-75% of speech at normal levels.<br>Requires: Bilateral hearing aids, FM systems for classroom | 800-1,500 |
| **Moderately severe**<br>(50-64 dB HL) | 3.83% | 207,914 | Misses 100% conversational speech without amplification.<br>Needs: Powerful hearing aids, speech therapy, educational support | 1,500-3,000 |
| **Severe**<br>(65-79 dB HL) | 3.29% | 178,667 | No speech perception without amplification.<br>May benefit from cochlear implant if aids insufficient | 3,000-35,000 |
| **Profound**<br>(80-94 dB HL) | 4.93% | 267,183 | Limited benefit from hearing aids alone.<br>Strong cochlear implant candidate; sign language often primary | 35,000-50,000 |
| **Complete**<br>(≥95 dB HL) | 0.38% | 20,687 | No auditory perception.<br>Requires: CI + intensive rehabilitation or sign language education | 50,000-70,000 |
| **INTELLECTUAL DISABILITY (n=2,560,545)** |  |  |  |  |
| **Borderline**<br>(IQ 70-84) | 13.38% | 342,526 | Slower learning pace; can achieve functional academics.<br>Support: Tutoring, modified curriculum, transition planning | 2,000-5,000/yr |
| **Mild**<br>(IQ 50-69) | 48.31% | 1,236,955 | 2-4 years behind in mental age; functional literacy possible.<br>Needs: Special education, some supervision | 5,000-10,000/yr |
| **Moderate**<br>(IQ 35-49) | 18.49% | 473,562 | Basic self-care; simple communication; supervised work possible.<br>Requires: Substantial support, specialized programs, group homes | 10,000-20,000/yr |

Table 2 – continued from previous page
| Condition & Severity Level | Percentage | Number | Functional Impact & Intervention | Cost (USD) |
| --- | --- | --- | --- | --- |
| **Severe**<br>(IQ 20-34) | 15.80% | 404,622 | Limited communication; requires intensive individualized support in all settings.<br>Full-time educational support: specialized curriculum, augmentative communication, life skills instruction | 20,000-40,000/yr |
| **Profound**<br>(IQ <20) | 4.02% | 102,879 | Minimal communication; requires intensive support for all daily activities.<br>Intensive support: 24-hour care, medical complexity common | 40,000-80,000/yr |

Cost ranges are expressed in 2023 US dollars. Percentages for each condition category sum to 100%. 327
*Abbreviations:* LAC = Latin America and Caribbean. dB HL = decibels Hearing Level. IQ = 328
Intelligence Quotient. CI = Cochlear Implant. VF = Visual Field. 329
*Sources:* Global Burden of Disease 2023; WHO International Classification of Functioning, Disability 330
and Health; Educational and rehabilitation cost estimates from regional health economic studies. 331

For vision loss, the high concentration in the moderate category (86.37%) highlights both the magnitude of the need and the opportunity for effective support. Approximately 1,121,609 children may experience barriers to learning associated with unaddressed refractive errors, many of whom could benefit from access to simple spectacles costing US$25–35. The smaller proportion requiring more specialized support (blindness 4.72%, severe 5.97%) necessitates targeted services but represents a manageable demand for regional capacity building (S3 Table).

Hearing loss severity distribution similarly favors intervention, with 87.57% of affected children (4,749,393 individuals) in mild-to-moderate categories where conventional hearing aids provide substantial benefit. However, the 5.31% (287,870 children) with profound or complete hearing loss potentially requiring cochlear implants represents a significant technological and financial challenge, with each implant costing US$35,000-50,000 plus lifetime support (S5 Table).

Intellectual disability presents a more complex distribution. While 61.69% fall into borderline-to-mild categories and may achieve functional independence with appropriate support, the 38.31% with moderate-to-profound disability require intensive, lifelong support systems that remain limited in most LAC countries.

### Comparison with OECD Countries: Identifying Gaps

Comparative analysis with OECD countries revealed striking epidemiological differences suggesting both challenges and opportunities (Table 1).

Hearing loss prevalence was 33.1% higher in LAC (3.63% vs 2.73%, p<0.001), with the excess concentrated in severe categories (50% higher prevalence of severe-to-complete hearing loss). This differential likely reflects higher exposure to risk factors including untreated otitis media (estimated 10-15% of LAC children vs 2-3% in OECD), ototoxic medication use without monitoring, environmental noise in unregulated urban settings, and limited neonatal hearing screening coverage (30% LAC vs 95% OECD).

Paradoxically, vision loss prevalence was 26.3% lower in LAC (0.87% vs 1.18%, p<0.001). This unexpected finding likely reflects measurement artifacts rather than genuinely better eye health. Possible explanations include: (1) higher OECD screening rates detecting mild presentations missed in LAC, (2) myopia epidemic in East Asian OECD countries not yet affecting LAC, (3) earlier correction in OECD preventing progression to measurable impairment, or (4) systematic under-diagnosis in LAC rural areas.

Intellectual disability showed 13.6% lower prevalence in LAC (1.71% vs 1.98%, p<0.001), almost certainly reflecting under-diagnosis rather than lower true prevalence. Given higher rates of risk factors in LAC (malnutrition, prematurity, perinatal complications), the expected prevalence would be equal or higher than OECD. The gap suggests 400,000-1,200,000 children with intellectual disability remain unidentified in LAC, with profound implications for service planning.

### Critical Intervention Windows: Timing Matters

Analysis of intervention timing revealed stark differences in outcomes based on developmental windows (Table 3).

**Table 3.** Comprehensive economic analysis of childhood disabilities in LAC: costs, investments, and returns.

| Economic Component | Vision | Hearing | Intellectual | Total |
| --- | --- | --- | --- | --- |
| ANNUAL COSTS (Billion USD) |  |  |  |  |
| Direct medical | 0.3-0.5 | 1.2-1.8 | 2.5-3.8 | 4.0-6.1 |
| Special education | 0.8-1.2 | 2.1-3.2 | 3.5-5.2 | 6.4-9.6 |
| Productivity loss | 1.5-2.2 | 3.2-4.5 | 2.0-3.0 | 6.7-9.7 |
| Family impact | 0.2-0.3 | 0.5-0.8 | 1.5-2.5 | 2.2-3.6 |
| Total cost | 2.8-4.2 | 7.0-10.3 | 9.5-14.5 | 19.3-29.0 |
| INTERVENTION INVESTMENT (Annual, Billion USD) |  |  |  |  |
| Screening | 0.05-0.08 | 0.15-0.25 | 0.10-0.15 | 0.30-0.48 |
| Devices/aids | 0.03-0.04 | 3.5-5.2 | N/A | 3.53-5.24 |
| Therapy | 0.10-0.15 | 0.8-1.2 | 4.5-6.8 | 5.4-8.15 |
| System | 0.02-0.03 | 0.05-0.08 | 0.15-0.25 | 0.22-0.36 |
| Total | 0.20-0.30 | 4.5-6.73 | 4.75-7.2 | 9.45-14.23 |
| COST-EFFECTIVENESS |  |  |  |  |
| \$/DALY averted | 15-40 | 1,850-2,950 | 5,000-15,000 | – |
| \$/case treated | 150-230 | 830-1,240 | 1,850-2,810 | – |
| CE status<sup>a</sup> | Very CE | CE | CE-marginal | – |
| ROI (20-year) |  |  |  |  |
| Benefit:cost | 10-15:1 | 5-8:1 | 2-3:1 | 4-6:1 |
| Payback (years) | 0.8-1.2 | 3.5-5.0 | 6-9 | 3.5-5.5 |
| Productivity gain (\$K) | 28 | 45 | 22 | – |
| Education gain (yr) | 2.1 | 1.8 | 0.9 | – |

All values in 2023 USD. Cost ranges reflect uncertainty in regional estimates. Human capital approach with 3% discount rate. <sup>a</sup>CE: <1×GDP/capita (\$9,276) = very cost-effective; <3×GDP/capita (\$27,828) = cost-effective. CE = Cost-effective; DALYs = Disability-Adjusted Life Years; ROI = Return on Investment. Sources: WHO-CHOICE database; World Bank national accounts; published economic evaluations from LAC countries.

The current economic costs associated with disability prevalence and its impacts are estimated at US$19.3-29.0 billion annually, equivalent to 0.34-0.51% of regional GDP. The largest component is associated with special education needs (US$6.4-9.6 billion), highlighting costs to education systems that may be reduced through early screening and support. Future productivity losses, calculated using age-earnings profiles and employment probability reductions, amount to US$6.7-9.7 billion annually for each affected cohort.

Investment requirements for comprehensive intervention programs total US$9.45-14.23 billion annually, approximately half the current cost of inaction. This investment would provide: universal screening programs at critical ages, devices and assistive technologies for all who need them, therapy and educational support services, health system strengthening including workforce training, and family support and education programs.

The economic case for intervention is compelling across all three conditions but varies in magnitude. Vision programs demonstrate exceptional returns (10-15:1 benefit-cost ratio) due to low intervention costs and high impact on educational and economic outcomes. A child receiving glasses at age 6 gains an average 2.1 years of education and US$28,000 in lifetime earnings for an investment of US$150-230.

Hearing interventions show strong returns (5-8:1) despite higher device costs. Early identification and hearing aid provision yields 1.8 additional years of schooling and US$45,000 in lifetime productivity gains. While cochlear implants are expensive (US$35,000-50,000), they remain cost-effective for profound hearing loss.

Intellectual disability interventions, while showing the lowest returns (2-3:1), remain economically justified. Early intervention programs costing US$5,000-10,000 annually can enable mild intellectual disability individuals to achieve supported employment, generating US$22,000 in lifetime productivity while reducing family burden and institutionalization costs.

### Health System Capacity and Service Gaps

Assessment of current service availability revealed substantial gaps between identified needs and service provision.

For vision loss, while 60-70% of affected children are eventually detected (often years late), only 25-35% receive and consistently use corrective lenses. Barriers include cost (even low-cost glasses exceed family means), availability (limited optical services in rural areas), cultural factors (stigma, cosmetic concerns), and quality (inappropriate prescriptions, poor-fitting frames).

Hearing loss shows the weakest cascade, with only 30% detected (usually after language delays manifest), 20% receiving proper audiological assessment, 10-15% obtaining hearing aids, and <5% of children with severe presentations accessing cochlear implants. The “lost to follow-up” rate between detection and intervention approaches 50%, reflecting fragmented referral systems.

Intellectual disability services are similarly inadequate, with 40% identified (often only moderate-severe presentations), 25% receiving comprehensive assessment, 20% accessing special education services, and <10% receiving evidence-based early intervention during the critical 0-3 year window.

## Discussion

This comprehensive analysis of sensory and intellectual disabilities among 149.3 million children and adolescents across 25 countries in Latin America and the Caribbean (LAC) highlights both the prevalence and distribution of these conditions and important opportunities to strengthen support systems. The estimated 9.3 million children represent not only a significant population but also an opportunity to enhance human potential through evidence-based policies and services.

### Principal Findings in Context

This comprehensive analysis of 9.3 million children and adolescents estimated with sensory and intellectual disabilities across Latin America and the Caribbean reveals both the magnitude of unmet need and the transformative potential of evidence-based interventions. The overall prevalence of 6.22% aligns with but refines previous global estimates, while uncovering critical regional specificities that challenge assumptions about disability in middle-income settings.

The predominance of hearing loss as the leading disability (3.63% prevalence) diverges from high-income country patterns and reflects LAC’s unique epidemiological profile [19]. The 33% excess compared to OECD countries cannot be dismissed as measurement artifact given the biological plausibility of higher risk exposure. Untreated otitis media, affecting 10-15% of LAC children compared to 2-3% in high-income settings, creates a preventable pathway to permanent hearing loss [4]. The widespread use of ototoxic aminoglycosides without audiological monitoring in resource-limited settings, environmental noise in rapidly expanding informal urban settlements, and persistent vaccine-preventable infections (rubella, measles, mumps) compound the burden [3].

The concentration of children in mild-to-moderate categories across all three conditions highlights important opportunities to strengthen early identification and support. The finding that 89% of vision loss can be effectively addressed with simple spectacles points to important gaps in service delivery, with over one million children experiencing barriers to learning due to unaddressed refractive errors, despite the availability of low-cost solutions (US$25 per pair of glasses). The evidence suggests that these gaps are driven by constraints in service organization, financing, and outreach, rather than by limitations in available technology. [5].

### Severity Distributions: Opportunities Hidden in Plain Sight

The severity distributions identified in this analysis provide a useful basis for prioritizing support. The high concentration of vision loss in moderate categories that can be effectively addressed with glasses (86.37%) highlights a key opportunity for early intervention. The estimated 1.12 million children with unaddressed refractive errors represent a substantial population at risk of barriers to learning. Without appropriate support, these children may experience difficulties in classroom participation, fall behind academically, and face increased risks of school dropout, with potential long-term implications for educational and socioeconomic outcomes. [20] (S3 Table).

Similarly, the 87.6% of children with hearing loss who could benefit from conventional hearing aids, alongside those who learning sign language, challenges narratives of inevitable linguistic isolation. Modern digital hearing aids, while costly at US$800-3,000 per pair, can restore functional hearing for the vast majority of affected children, just as early exposure to sign language provides a rich and complete fondation for communication. The tragedy is not the technology gap but the coverage and accessibility gap—with current hearing aid coverage in LAC estimated at only 10-20% and limited acess to sign language education, millions of children remain trapped in preventable communication barriers [19].

The intellectual disability distribution, with 61.7% in borderline-to-mild categories, similarly challenges fatalistic approaches. These 1.58 million children are capable of meaningful participation, functional literacy, vocational skills, and semi-independent living with appropriate support. The stark reality that less than 30% access any special education services, and fewer than 10% receive early intervention during the critical 0-3 year window, represents a massive failure to realize human potential [21].

### The Paradox of LAC versus OECD Comparisons

The paradoxical findings of lower vision loss and intellectual disability prevalence in LAC compared to OECD countries merit careful interpretation. For vision loss, the 26% lower prevalence almost certainly reflects measurement and health system artifacts rather than genuinely better eye health. Several mechanisms likely contribute: (1) OECD’s universal vision screening detects mild presentations missed in LAC; (2) the myopia epidemic affecting East Asian OECD countries has not yet reached LAC; (3) earlier correction in OECD prevents progression to measurable impairment; and (4) systematic under-detection in LAC rural and indigenous populations where access to eye care is minimal [22].

The intellectual disability prevalence deficit is even more certainly an artifact of under-diagnosis. Given LAC’s higher rates of virtually every known risk factor—malnutrition (11% stunting), prematurity (8.5% of births), perinatal complications (1.5-3 per 1,000 births with HIE), environmental toxins, and limited early stimulation—the true prevalence should equal or exceed OECD levels [6]. Our estimate that 400,000-1,200,000 children with intellectual disability remain unidentified has profound implications for service planning. When these children eventually surface in school systems, they arrive without the benefit of early intervention, with capabilities far below their potential.

### Critical Windows: The Neuroscience of Lost Opportunity

The critical intervention windows identified in our analysis are not administrative conveniences but reflect fundamental neurodevelopmental processes. For hearing, the auditory cortex undergoes experience-dependent organization in the first year of life. Auditory deprivation during this period leads to cross-modal reorganization, with visual processing colonizing auditory regions, making later hearing restoration less effective [23]. Children identified and aided before 6 months show near-normal language trajectories, while those identified after 12 months show persistent deficits even with optimal intervention. Every month of delay during this critical period translates to measurable linguistic disadvantage persisting into adulthood.

For vision, while the critical period for binocular vision closes by age 7-8, the window for educational impact extends through school entry. Children who receive glasses at ages 6–7 can improve their academic trajectories within 1–2 years, whereas those without access to appropriate support until adolescence may experience more persistent learning gaps. The underlying mechanisms extend beyond visual clarity to include cumulative missed learning opportunities, as early difficulties in accessing classroom materials can affect foundational skills such as reading and influence subsequent learning [24].

Intellectual disability presents the widest but earliest closing window. The 0-3 period of synaptic proliferation and pruning represents unparalleled neuroplasticity. High-quality early intervention during this period can shift developmental trajectories by 0.7-1.2 standard deviations, equivalent to 10-18 IQ points. After age 5, the same intervention intensity yields minimal cognitive gains, though adaptive behavior can still improve. This is not therapeutic nihilism for older children but recognition that earlier intervention yields exponentially greater returns [21].

### Economic Arguments: Beyond Humanitarian Imperatives

While humanitarian arguments for addressing childhood disabilities are compelling, the economic case provides the pragmatic foundation for political action. Our analysis reveals that the current cost of inaction (US$19.3-29.0 billion annually) exceeds the investment needed for comprehensive intervention (US$9.45-14.23 billion) by a factor of two. This is before considering the intangible costs of impact on families and caregivers, social exclusion, and unrealized human potential.

The variation in cost-effectiveness across conditions provides guidance for staged implementation. Vision programs, with costs of US$15-40 per DALY averted, rank among the most cost-effective health interventions available, comparable to vaccination programs. The 10-15:1 benefit-cost ratio means that every dollar invested in school vision screening and glass provision returns US$10-15 in economic benefits—few investments in any sector offer such returns [25].

Hearing interventions, while more expensive at US$1,850-2,950 per DALY averted, remain highly cost-effective by WHO standards (<3*×* GDP per capita). The 5-8:1 benefit-cost ratio, while lower than vision programs, still represents exceptional returns. The higher costs reflect device expenses, but technological advances including 3D-printed hearing aids and tele-audiology are reducing costs while maintaining quality [19].

Intellectual disability interventions present the highest costs but remain economically justified. The 2–3:1 benefit–cost ratio indicates positive returns. Moreover, the benefits extend beyond the individual to families and caregivers, contributing to reduced support needs and broader social and economic gains. [26].

### Financing the Intervention: Securing Educational and Economic Futures

The economic imperative of these interventions fundamentally rests on protecting the human capital of school-aged children. Failing to provide timely screening, assistive technologies, and specialized support directly compromises their educational attainment and future income-generating capacity, perpetuating intergenerational cycles of poverty and inequality. Therefore, funding these interventions must be recognized not as a discretionary health expenditure, but as a critical investment to secure the region’s future workforce and macroeconomic stability.

To bridge the estimated US$9.45-14.23 billion funding gap in a fiscally constrained region, LAC governments must diversify their financing strategies beyond traditional health budgets. Viable alternatives include the creation of dedicated national or regional funds for early childhood development and disability inclusion. These funds can be sustainably capitalized through earmarked taxation on specific sectors, such as health taxes on harmful products (e.g., tobacco, alcohol, or ultra-processed foods). Furthermore, governments must actively leverage international financing organizations and multilateral development banks to secure the upfront investments required, ensuring that no child’s educational and economic potential is forfeited due to a lack of immediate domestic fiscal space.

### Implementation Framework: From Evidence to Action

Based on our findings, we propose a three-tiered implementation framework that recognizes both the heterogeneity of country capacities and the urgency of action:

### Tier 1 - Universal Quick Wins (Implementation: 0-2 years)

Vision screening at school entry with immediate glass provision represents the highest-impact, lowest-cost intervention. Countries could achieve 80% coverage within 2 years using: teachers trained in basic vision screening (2-day training), partnerships with optical suppliers for bulk purchasing (reducing unit costs to US$10-15), mobile units for rural areas, and recycling programs for frame reuse. Chile’s experience demonstrates feasibility, achieving 85% coverage within 18 months of program launch [27].

Basic hearing screening in schools using tablet-based audiometry could identify the bulk of undetected children. While not replacing newborn screening, school-entry screening captures those missed earlier and children with acquired hearing loss. Costa Rica’s program shows 90% sensitivity using minimally trained personnel [28].

Developmental screening at well-child visits using validated tools (ASQ, Denver II) could identify intellectual disability during the critical window. Brazil’s integration into primary care demonstrates feasibility even in resource-limited settings [29].

### Tier 2 - Systematic Capacity Building (Implementation: 2-5 years)

Establishing newborn hearing screening requires greater infrastructure but remains achievable through phased implementation: starting in tertiary hospitals (covering 40-50% of births), expanding to secondary facilities, and eventually reaching primary level. The 1-3-6 protocol (screening by 1 month, diagnosis by 3 months, intervention by 6 months) should guide implementation [30].

Strengthening special education services through teacher training, curriculum adaptation, and resource provision requires sustained investment but builds on existing educational infrastructure. Colombia’s inclusive education program provides a replicable model [31].

Developing early intervention programs for intellectual disability using cascading training models can rapidly expand coverage. Parent-mediated interventions show particular promise in resource-limited settings [32].

### Tier 3 - Comprehensive Systems (Implementation: 5-10 years)

Universal coverage of complex interventions (cochlear implants, intensive behavioral support) requires substantial infrastructure development but should remain the ultimate goal. Regional cooperation could facilitate bulk purchasing, training programs, and technical assistance.

Integrated service delivery models that address multiple disabilities through single entry points reduce fragmentation and improve outcomes. Mexico’s CRIT centers demonstrate the efficiency of comprehensive rehabilitation services [33].

### Implications for Policy and Practice

Our findings have immediate implications for policy formulation and resource allocation:

#### Act on opportunities for inclusive support

The predominance of mild-to-moderate and often treatable levels highlights important opportunities for early identification and support. These findings suggest the need to strengthen service delivery and inclusive systems. Children with disabilities should be recognized not only as recipients of services but as active participants whose inclusion in educational and social systems contributes to broader societal outcomes.

#### Prioritize early identification

Every month of delay during critical periods can lead to lasting impacts on development and learning. Screening programs must be treated as essential services, not optional additions.

#### Integrate across sectors

Disability sits at the intersection of health, education, and social protection. Siloed approaches perpetuate inefficiency and gaps.

#### Address stigma systematically

Cultural barriers can be as significant as, or in some contexts exceed, economic barriers. Public education campaigns, including those that highlight diverse experiences of persons with disabilities, can contribute to shifting perceptions.

#### Leverage digital health innovations

Tele-audiology, smartphone-based vision screening, and validated app-based developmental assessments can overcome geographic barriers and exponentially expand diagnostic reach at a minimal marginal cost.

#### Build regional cooperation

Small countries cannot achieve economies of scale alone. Regional purchasing agreements, training programs, and technical standards benefit all.

#### Monitor and evaluate rigorously

Investment without measurement wastes resources. Simple indicators (screening coverage, device provision, educational outcomes) should guide program refinement.

### Strengths and Limitations

This study’s strengths include comprehensive coverage of 25 countries representing >95% of LAC’s population, use of standardized GBD methodology enabling valid comparisons, integration of epidemiological and economic analyses providing policy-relevant insights, severity stratification moving beyond simple prevalence to intervention planning, and synthesis of critical windows evidence guiding program timing.

Several limitations warrant consideration. First, GBD estimates rely on modeled data where primary sources are unavailable, potentially masking subnational variations particularly important in countries with substantial indigenous populations or geographic barriers. Second, disability definitions and severity thresholds may not fully capture cultural variations in functioning across diverse LAC contexts. Third, economic impact calculations necessarily simplify complex cascading effects of childhood disability on families, communities, and intergenerational mobility. The cross-sectional design prevents causal inference about risk factors or intervention effectiveness.

Under-diagnosis, particularly in rural and indigenous populations, likely leads to systematic underestimation. The exclusion of other important disabilities (autism spectrum disorder, cerebral palsy, multiple disabilities) underestimates total burden. Economic projections involve uncertainty about future labor markets, technological costs, and implementation efficiency. The assumption of linear relationships between intervention timing and outcomes may oversimplify complex developmental processes. Finally, this study employs biomedical prevalence data from the GBD framework, which does not capture the environmental, attitudinal, and policy barriers that, according to the social model of disability enshrined in the UN Convention on the Rights of Persons with Disabilities (CRPD), are central to the experience of disability. Our findings should be interpreted within this limitation.

### Future Research Priorities

Critical knowledge gaps requiring urgent research include:

#### Implementation science

How can evidence-based interventions be adapted for diverse LAC contexts while maintaining fidelity? What implementation strategies maximize coverage and quality?

#### Diagnostic validation

Current screening tools require validation in LAC populations, particularly indigenous groups with different backgrounds and cultural expressions of disability.

#### Longitudinal outcomes

Cohort studies tracking children from early intervention through adulthood would provide LAC-specific evidence on long-term impacts.

#### Economic evaluation

Prospective economic evaluations of intervention programs would refine cost-effectiveness estimates and guide resource allocation.

#### Intersectionality

How do multiple disadvantages (disability + poverty + indigenous status + rural residence) interact? What additional support do multiply disadvantaged children require?

#### Technology assessment

Rigorous evaluation of emerging technologies (3D-printed devices, AI-based screening, tele-rehabilitation) could identify cost-effective innovations.

#### Family impacts

Better understanding of disability’s effects on siblings, parental employment, and family stability would strengthen the case for family-centered interventions.

## Conclusions

The 9.3 million children and adolescents estimated with sensory and intellectual disabilities in Latin America and the Caribbean represent both a moral imperative and economic opportunity demanding immediate action. Our comprehensive analysis reveals that far from being an intractable burden, the majority of these disabilities are either effectively addressed (vision), treatable (hearing), or manageable (intellectual) with established, cost-effective interventions. The tragic reality is not technological inadequacy but systemic failure to implement proven solutions.

The concentration of estimated children in mild-to-moderate severity categories amenable to intervention fundamentally reframes the disability challenge. Given that 89% of vision loss can be effectively addressed with simple spectacles, 87.6% of hearing loss responds to conventional hearing aids, and 61.7% of children with intellectual disability may achieve substantial autonomy with appropriate support, these findings highlight important opportunities to strengthen early and sustained support systems.

The economic argument is equally compelling. With current costs of inaction exceeding required investments by a factor of two, and positive returns on investment across all three conditions, addressing childhood disabilities is not just morally right but economically rational. The exceptional cost-effectiveness of vision programs (US$15-40 per DALY averted) places them among the highest-value health interventions available, while even the most expensive intellectual disability interventions generate positive economic returns.

Critical intervention windows in early life underscore the urgency of action. For hearing, the 1-3-6 protocol must become standard practice, with every month of delay during the first year translating to measurable, persistent linguistic disadvantage. For vision, school-entry screening with immediate correction can eliminate academic achievement gaps within two years. For intellectual disability, intensive intervention during the 0-3 neuroplasticity window can shift developmental trajectories by 10-18 IQ points, while the same resources applied later yield minimal cognitive gains.

The path forward requires not technological innovation but political will and systematic implementation. Countries like Costa Rica, Chile, and Cuba demonstrate that comprehensive disability programs are achievable even with limited resources when political commitment exists. The proposed three-tiered implementation framework—starting with universal quick wins, building systematic capacity, and ultimately achieving comprehensive coverage—provides a pragmatic roadmap respecting diverse country capacities while maintaining urgency.

Regional cooperation through existing institutions (PAHO, IDB, ECLAC) can accelerate progress through bulk purchasing, shared training programs, and technical assistance. The recent establishment of regional disability frameworks provides institutional support, but implementation has lagged. Our findings provide the evidence base to move from aspirational declarations to measurable action.

The implications extend beyond the affected children to families, communities, and societies. Every child whose vision is corrected stays in school longer, earns more, and contributes more to society. Every child who receives timely hearing intervention develops language, forms relationships, and participates fully in community life. Every child with intellectual disability who receives early support achieves greater independence, reducing family burden and institutional costs.

These findings suggest that the fiscal debate facing Latin America and the Caribbean is shifting from whether action is affordable to how investments can be sequenced to minimise avoidable losses. Each year in which interventions are deferred sustains preventable limitations for hundreds of thousands of children, with downstream consequences for poverty and productivity. The evidence assembled here delineates feasible solutions and associated economic returns, offering governments a concrete basis for prioritizing implementation. Achieving impact will require translating knowledge into action through coordinated policies and financing across health, education, and social protection systems.

As we look toward 2030 and the Sustainable Development Goals, addressing childhood disabilities emerges not as one priority among many but as estimated foundational to achieving inclusive, equitable development. The 9.3 million affected children cannot wait for perfect solutions or ideal conditions. Clearly planned interventions implemented now could transform millions of lives while building systems for future comprehensive coverage.

The effectiveness of current responses to childhood disability will ultimately be reflected not only in policies and frameworks, but in the extent to which children are supported to participate fully in educational and social systems. The analyses presented here narrow the room for inaction by clarifying the opportunity costs of delay. Progress will depend on sustained commitment, intersectoral coordination, and accountability mechanisms that ensure no child in Latin America and the Caribbean is left without access to appropriate support and services.

## Acknowledgments

We thank the Institute for Health Metrics and Evaluation for providing access to GBD 2023 data, the Inter-American Development Bank for supporting this analysis, WHO/PAHO for technical guidance, national statistical offices and health ministries for primary data, and the disability community for insights into lived experiences. Special recognition to the 9.3 million estimated children and adolescents living with disabilities in LAC, whose potential drives our commitment to evidence-based action.

## Author contributions

**J.A.P.M.C.:** Conceptualization, Methodology, Data curation, Formal analysis, Validation, Visualization, Writing–original draft, Writing–review & editing.

**E.N.H.:** Conceptualization, Visualization, Supervision, Writing–original draft, Writing–review & editing.

**B.G.A.:** Conceptualization, Methodology, Data curation, Validation, Visualization, Writing–original draft, Writing–review & editing.

**A.N.P.:** Methodology, Validation, Formal analysis, Writing–review & editing.

## Data Availability

All data were obtained from publicly available sources. Global Burden of Disease 2023 estimates are accessible through the Institute for Health Metrics and Evaluation Global Health Data Exchange (https://ghdx.healthdata.org/gbd-results-tool).

## Supporting Information S1: ROI Assumptions and Input Parameters

This supplementary table documents the economic assumptions underlying the return on investment (ROI) calculations presented in the main text (Table 3).

**Table 4.**
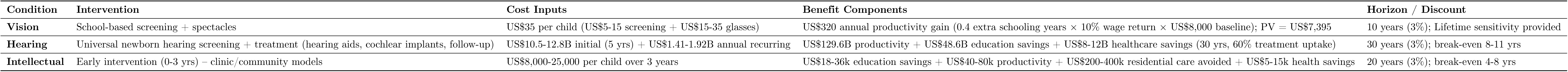
ROI Input Parameters and Assumptions by Condition.

### Data Sources

- Vision: IDB policy brief on vision loss and blindness in school-age children in LAC (2025)
- Hearing: IDB policy brief on hearing loss and deafness in school-age children in LAC (2025)
- Intellectual: IDB analytical inputs on intellectual disability in school-age children in LAC (forthcoming)

### Notes

All ROI calculations use a 3% annual discount rate following WHO-CHOICE guidelines. Sensitivity analyses varying cost inputs, effectiveness parameters, and discount rates confirm robustness of cost-effectiveness conclusions across scenarios.

